# Mechanisms linking physical activity with psychiatric symptoms: a protocol for a systematic review

**DOI:** 10.1101/2022.01.19.22269541

**Authors:** Phuong Thuy Nguyen Ho, Tram Ha Pham Bich, Thao Tong, Wichor M Bramer, Amy Hofman, David R Lubans, Meike W Vernooij, María Rodriguez-Ayllon

## Abstract

**Introduction:** Persistent psychiatric symptomatology during childhood and adolescence predicts vulnerability to experience mental illness in adulthood. Physical activity is well-known to provide mental health benefits across the lifespan. However, the underlying mechanisms linking physical activity and psychiatric symptoms remain underexplored. In this context, we aim to systematically synthesize evidence focused on the mechanisms through which physical activity might reduce psychiatric symptoms across all ages.

**Methods and analysis:** With the aid of a biomedical information specialist, we will develop a systematic search strategy based on the predetermined research question in the following electronic databases: MEDLINE, Embase, Web of Science, Cochrane, and PsycINFO. Two independent reviewers will screen and select studies, extract data, and assess the risk of bias. In case of inability to reach a consensus, a third person will be consulted. We will not apply any language restriction, and we will perform a qualitative synthesis of our findings as we anticipate that studies are scarce and heterogeneous.

**Ethics and dissemination:** Only data that has already been published will be included. Then, ethical approval is not required. Findings will be published in a peer-reviewed journal and presented at conferences. Additionally, we will communicate our findings to healthcare providers and other sections of society (e.g., through regular channels, including social media).

**PROSPERO registration number:** CRD42021239440

**Strengths and limitations of this study:**

- This protocol has been designed according to the Preferred Reporting Items for Systematic Reviews and Meta Analyses for Protocols (PRISMA-P) guidelines and guidelines of the Cochrane Effective Practice and Organisation of Care.
- This protocol presents a cautiously designed search strategy, inclusion and exclusion criteria, and timespan and age-range coverage.
- A possible limitation is that included studies might be heterogeneous in the study design, data collection methods, and data analysis which might limit the ability to synthesize the results using a meta-analysis.
- The value of this systematic review depends on the quality and availability of the evidence on the topic.

## INTRODUCTION

Persistent psychiatric symptomatology in childhood and adolescence predicts vulnerability to experience mental illness later in life^1^. Specifically, individuals with mental illness have a decreased life expectancy of 10–15 years^2^ and a lower quality of life^3^ than individuals from the general population. Psychiatric symptoms are typically grouped into two broad categories (i.e., internalizing/emotional, and externalizing/behavioral)^4^. Specifically, the externalizing problems include a variety of disinhibited/externally-focused behavioral symptoms such as conduct problems, rule-breaking behavior, attention-deficit/hyperactivity problems. On the contrary, the internalizing disorder include a variety of over-inhibited/internally-focused symptoms, such as depression, anxiety, or somatic symptoms. Several risk factors for psychiatric symptoms have been well established in childhood (e.g., poverty and social disadvantage)^5^ and adulthood (e.g., level of education and physical illness)^6^. However, less is known about the protective factors (e.g., physical activity) that might contribute to decreasing both child and adult psychopathology.

Physical activity is well-known to provide multiple health-related benefits across the lifespan^7^. In particular, there is a growing body of literature suggesting that physical activity has a small-to-moderate effect on psychiatric symptoms in childhood and adolescence^5,6,7^ but also in adulthood^11,12^. However, most of the studies have focused on exploring the effect size of the association or effect in terms of dose-response, while the mechanisms underlying this relationship or effect remain underexplored. In 2016, Lubans et al.^13^ published a systematic review of the mechanisms linking physical activity and psychiatric symptoms in children and adolescents. They proposed a conceptual model, which postulated three distinct yet intertwined potential groups of mechanisms (i.e., neurobiological, psychosocial, and behavioral mechanisms). In brief, they identified a lack of available evidence for the specific mechanisms responsible for the effect of physical activity on mental and cognitive health in young people. Additionally, none of the studies included in their review examined potential mechanisms responsible for the effects of physical activity on mental health in young people using an accepted statistical analysis (e.g., statistical mediation analysis)^11^. Lastly, they only included intervention studies, and although this type of design can provide evidence for cause and effect, observational studies can also provide complementary information, particularly when there is a lack of evidence on the topic.

In adults, only narrative reviews^12,13,14^, mainly focused on cognition^12^ and depression^13,14^, have explored the potential mechanisms that might link physical activity with psychiatric symptoms in adulthood. For instance, Stillman et al.^12^ suggested that physical activity might reduce depression and anxiety via psychosocial pathways (e.g., mood). Additionally, Kandola et al.^16^ presented a conceptual framework of the key biological and psychosocial mechanisms underlying the relationship between physical activity and depressive symptoms in adults. However, no previous systematic reviews have been performed to synthesize the existing evidence in adults.

Understanding the mechanisms linking physical activity with psychiatric symptoms may help to explain, predict, and intervene more effectively, which could stimulate the identification of cost-efficient alternative therapies for preventing and treating mental illness at all ages. To establish this evidence-based, it is imperative to synthesize and update all relevant literature mapping the mechanisms through which physical activity reduces psychiatric symptoms across the lifespan.

### Objective

We aim to conduct a systematic review to explore the underlying mechanisms linking physical activity with psychiatric symptoms in humans of all ages.

### Review questions

How does physical activity affect/associate with psychiatric symptoms via psychosocial, neurobiological, and behavioral pathways across the lifespan?

## METHODS

The present protocol follows the PRISMA-P guideline for systematic review and meta-analysis protocols^18^.

### Patient and public involvement

Patients and the public were not involved in the design, development, conduct, reporting or dissemination of this study.

### Eligibility criteria

We will include studies based on predefined criteria as summarized in **Table 1** and the text below^19^.

**Table 1.** Inclusion criteria based on PICOS strategy.

| **PICOS** | **Inclusion criteria** | **Exclusion criteria** |
| --- | --- | --- |
| **Population** | 1. All ages across the lifespan: infancy and toddlerhood (birth to age 2), preschoolers (2–5 years), children (6–11 years), adolescents (12–18 years), young and middle adults (18– 65), late adulthood (+65).<br>2. Human studies. | 1. Studies including individuals with physical or psychological disorders diagnosed by medical records.<br>2. Elite athletes.<br>3. Animal studies. |
| **Intervention** | 1. Observational studies, which explored the mechanisms through which physical activity is associated with psychiatry symptoms.<br>2. Studies examining the mechanisms through which physical activity has a positive effect on psychiatry symptoms. | 1. Multiple health behavior intervention studies (e.g., co-interventions such as a dietary program combined with physical activity).<br>2. Studies in which physical fitness (i.e., capacity to perform physical activity, which refers to a full range of physiological and psychological qualities)<sup>20</sup>, or sedentary behavior (i.e., any waking behavior characterized by an energy expenditure ≤ 1.5 METs, while in a sitting, reclining or lying posture)<sup>21</sup> are the independent variables instead of physical activity (i.e., any bodily movement produced by skeletal muscle that results in energy expenditure)<sup>22</sup>. |
| **Comparison** | 1. Not applicable |  |
| **Outcomes** | 1. The subscales of internalizing symptoms (i.e., depression, anxiety, somatic symptoms) and externalizing symptoms (i.e., conduct problems, rule-breaking behavior, attention deficit/hyperactivity problems). |  |
| **Study design** | 1. Intervention studies (randomized controlled trials, non-randomized control trials), prospective longitudinal studies and cross-sectional studies. | 1. Conference proceedings and other types of grey literature.<br>2. Narrative reviews, systematic reviews, or meta-analyses. |

#### Population

We will include human studies including participants of all ages. Studies including individuals with physical or psychological disorders diagnosed by medical records, elite athletes, and animals will be excluded.

#### Intervention

We will include any observational studies, which have explored the mechanisms through which physical activity is associated with psychiatric symptoms. Intervention studies examining the mechanisms through which physical activity affects psychiatric symptoms will be also included. Studies in which physical fitness (i.e., capacity to perform physical activity, which refers to a full range of physiological and psychological qualities)^20^, or sedentary behavior (i.e., any waking behavior characterized by an energy expenditure ≤ 1.5 METs, while in a sitting, reclining or lying posture)^21^ are the independent variables instead of physical activity (i.e., any bodily movement produced by skeletal muscle that results in energy expenditure)^22^ will be excluded. Additionally, multiple health behavior intervention studies (e.g., co-interventions such as a dietary program combined with physical activity) will be excluded because they preclude drawing conclusions on the isolated effect of physical activity or sedentary behavior on psychiatric symptoms.

#### Outcomes

We will include the subscales of internalizing (i.e., depression, anxiety, somatic symptoms) and externalizing (i.e., conduct problems, rule-breaking behavior, attention deficit/hyperactivity problems) disorders.

#### Study designs

Intervention studies (randomized controlled trials [RCT], non-RCTs), prospective longitudinal and cross-sectional studies will be included. We will not include conference proceedings and other types of grey literature since risk of bias for these studies cannot be adequately assessed^23^.

#### Potential mechanisms

Studies will be included if they explored the role of any potential neurobiological, psychosocial, and behavioral mechanisms in the relationship between physical activity and psychiatry symptoms.

#### Further restrictions

No language and publication date restriction will be applied. All databases will be searched from their date of inception, and we will include every study that meets the above-mentioned criteria regardless of the language.

### Search strategy for identifying relevant studies

With the assistance of a biomedical information specialist, we will develop a systematic search strategy based on the predetermined research question in the following electronic databases: MEDLINE Ovid, Embase.com, Web of Science Core Collection, Cochrane CENTRAL register of Trials, and PsycINFO Ovid. First, we will search for potentially relevant studies based on a search strategy that is the combination of Medical Subject Headings (MeSH) terms for Medline and Emtree terms for Embase and free text search. Our research team, including a librarian who is specialized in search strategy development, has developed this search strategy. Search terms are personalized to each database (see **Online supplemental appendix**). Search terms include four parts: (1) terms to identify our independent variable (i.e., physical activity); (2) terms to identify our mediating variables (i.e., neurobiological, psychosocial, behavioral mechanisms); (3) terms to identify our outcome (i.e., psychiatric symptoms); and (4) terms to exclude articles that match our exclusion criteria. An additional search for studies will be performed by screening reference lists of included studies and their citations through Google Scholar. Third, we will contact experts in the field to identify additional studies that may have been missed and any relevant ongoing or unpublished studies.

### Study records

#### Data management

First, we will extract all studies identified by the different sources into an EndNote Library. Second, we will use a published method that uses this software to automatically eliminate the duplicate studies^24^. In our final report, we will note the number of duplicates in the PRISMA flow diagram (see **Figure 1**).

**Figure 1.**
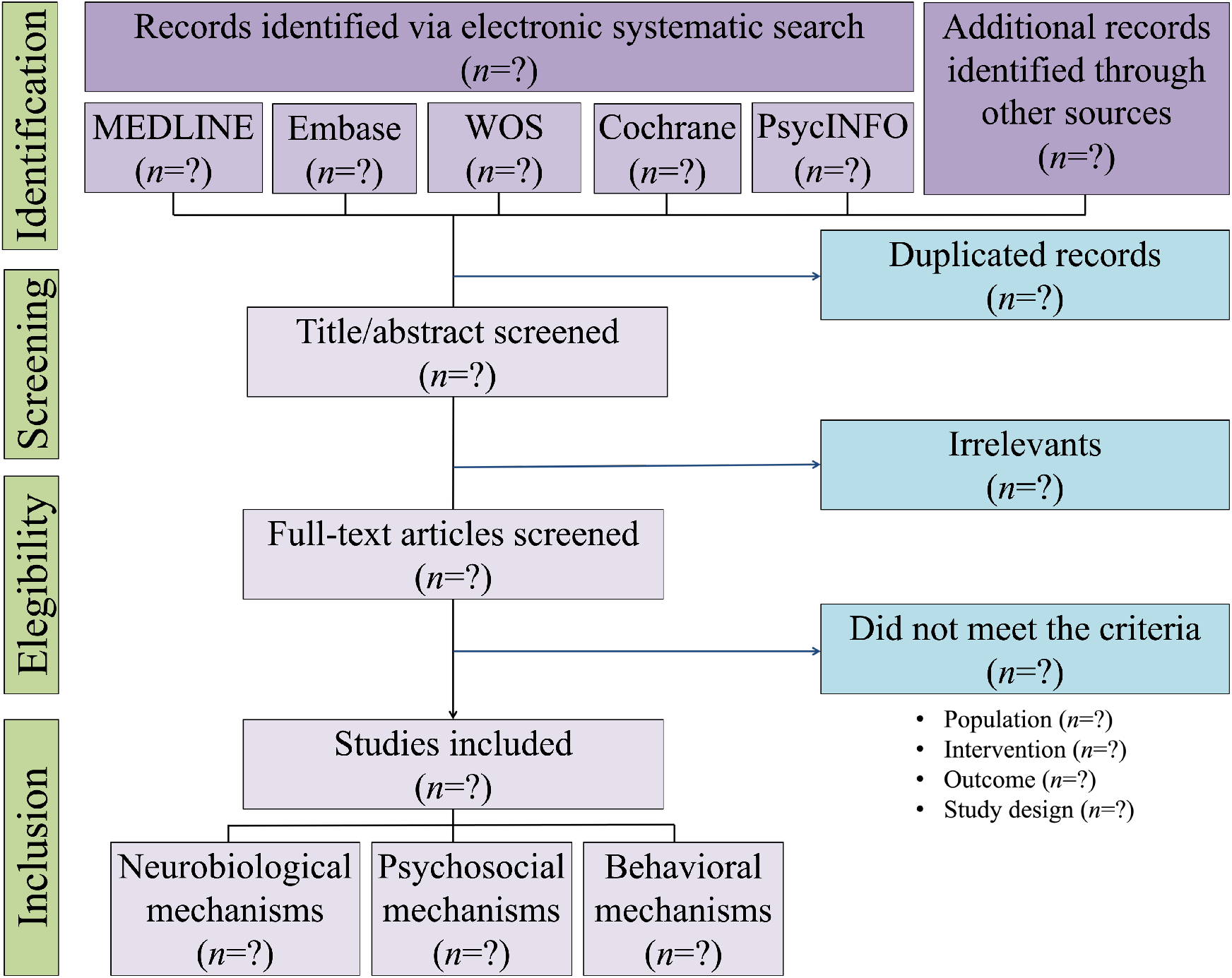
Flow diagram for study selection.

#### Selection process

First, two independent researchers (PTNH and THPB) will screen titles and the abstracts for eligibility. When disagreements emerge between the two independent researchers, consensus will be obtained through discussion or when required, the opinion of a third researcher (MR-A) will be considered. Second, we will then obtain the full-text reports of studies that may fit eligibility criteria based on this assessment. Afterward, the same two independent researchers (PTNH and THPB) will assess eligibility based on the full texts. Any discrepancies will be again resolved after discussion with a third researcher (MR-A).

### Data extraction process

Two researchers (PTNH and TT) will independently extract data from the included studies to a customized data extraction form developed a priori that has been piloted using one eligible study (see **Table 2**). Again, any discrepancies will be resolved after discussion with a third researcher (MR-A). We will contact authors for any relevant missing data.

**Table 2.**
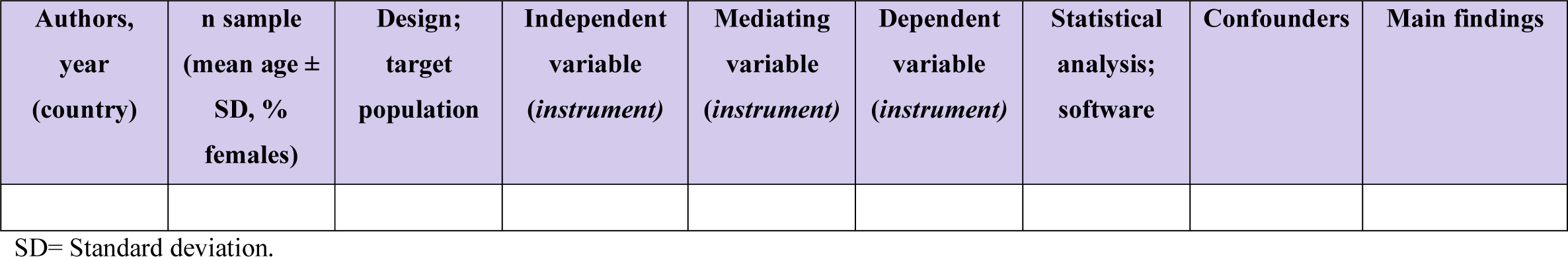
Summary of research investigating the mechanisms linking physical activity with psychiatric symptoms (n = ?).

From eligible studies, we will extract the following items: study background (name of the first author, year, and study location), sample characteristics (number of participants, age of participants, and percentage of female participants), design (intervention [RCT or non-RCT], or observational [cross-sectional or longitudinal]), independent variables (instruments), dependent variables (instrument), mediating variables (instrument), statistical analyses and software, confounders, and main findings. For intervention studies (RCTs and non-RCTs), we also extract weeks of intervention, description of the program, intensity, duration, and frequency. For longitudinal studies, we also extract years of follow-up.

### Risk of bias and quality of the evidence

The risk of bias will be evaluated independently by two researchers (PTNH and TT) and disagreements were solved in a consensus meeting with the same third researcher (MR-A). The risk of bias will be evaluated using the Joanna Briggs Institute Critical Appraisal Tool for Systematic Reviews (https://jbi.global/critical-appraisal-tools). This tool has already been used by other authors in the field^25,26^. In brief, this tool includes four specific checklists depending on the study design (i.e., cross-sectional studies, longitudinal studies, RCTs and non-RCT). There are four possible answers for each category: “yes” (criterion met), “no” (criterion not met), “unclear” or “not applicable”. The specific tools include: eight items for cross-sectional studies, 11 items for longitudinal studies, nine items for non-RCTs and thirteen items for RCTs. Studies will be categorized as “high risk” or “low risk”. Specifically, the studies will be considered as “low risk” if at least 75% of the applicable items are scored as “yes” (criterion met). In contrast, articles will be considered “high risk” when less than 75% of the applicable items were scored as “yes”. This classification has been previously employed by Molina-Garcia et al. ^27^.

Lastly, the Grading of Recommendations Assessment, Development and Evaluation framework will be used to assess the quality of the evidence across studies.

### Data synthesis and analysis

In case overlapping populations are analyzed in multiple studies, we will include according to the following hierarchy the study that (1) has the lowest risk of bias, or (2) incorporates the largest sample size. In the case when a study reports multiple effect estimates for overlapping populations, we will select according to the following hierarchy: (1) the most adjusted model, (2) the closest time-point to the end of the intervention, or (3) the largest treatment group. Findings from observational and intervention studies will be rated using the method first employed by Sallis et al.^25^, and more recently by Lubans et al.^13^, and Rodriguez-Ayllon et al.^7^. If 0–33% of studies reported a statistically significant mediation (e.g., self-esteem) between the independent (e.g., physical activity) and dependent variable (e.g., depressive symptoms), the result will be classified as no association (Ø); if 34–59% of studies reported a significant mediation, or if fewer than four studies reported on the outcome, the result will be classified as being inconsistent/uncertain (?); and if ≥ 60% of studies found a statistically significant mediation, the result will be classified as significant (✓).

## Data Availability

As systematic reviews use publicly available data, no formal ethical review and approval are needed.

## Ethics and dissemination

We will communicate our findings to researchers, pediatricians, health professionals, and lectures through scientific seminars and conferences. Additionally, we will disseminate our results using different approaches. Specifically, we will publish press articles in public journals and magazines, do radio and television interviews, and publish our findings in a scientific peer-review journal. We will also present our main results to policymakers and healthcare providers, which might impact policy and healthcare practice.

## Disclaimer

The funders of the present study did not have any role in the design, decision to publish or preparation of the protocol.

## Patient consent

Not required.

## Provenance and peer review

Not commissioned, externally peer reviewed.

## Open access

This is an open access article distributed in accordance with the Creative Commons Attribution Non-Commercial (CC BY-NC 4.0) license, which permits others to distribute, remix, adapt, build upon this work non-commercially, and license their derivative works on different terms, provided the original work is properly cited, appropriate credit is given, any changes made indicated, and the use is non-commercial. See: http://creativecommons.org/licenses/by-nc/4.0/.

## Authors’ contributions

MR-A, PTNH designed and drafted the protocol. WMB performed the search strategy. MR-A, PTNH, THPB, TT, AH, DRL, MV revised and approved the final version of the manuscript. MR-A will be the guarantor of the review.

